# Multifactorial tuberculosis severity score for people living with HIV based on the Rand Appropriateness Method

**DOI:** 10.64898/2026.06.21.26356185

**Authors:** Robert Akpata, Didier Laureillard, Cossi Angelo Attinsounon, Nathalie De Castro, Gisèle Badoum, Nicolas Veziris, Eugène Messou, François-Xavier Blanc, Laurence Weiss, Xavier Anglaret, Marcel Zannou, Olivier Marcy

## Abstract

**Background:** In people living with Human Immunodeficiency Virus (PLWH), tuberculosis (TB) is the leading cause of death and is often associated with substantial morbidity. Better identifying PLWH with severe forms of TB could help target early interventions to reduce mortality and severe morbidity. Existing TB severity assessment tools may be sub-optimal for assessing disease severity in PLWH, since they incompletely integrate key determinants of disease severity. We aimed to develop a consensus-based TB severity score tailored to PLWH.

**Methods:** We developed a multifactorial TB severity score (TBSS) for PLWH using a modified Delphi process with a multidisciplinary group of international TB experts as the second part of a RAND/UCLA Appropriateness Method, following a previously published systematic review.

**Results:** Eight of 15 invited experts (53%) participated in both Delphi rounds. Of 62 candidate factors, 15 reflecting TB-related characteristics, host-related characteristics as well as characteristics related to both TB and host were rated as having high appropriateness for inclusion in the final TBSS. The total score ranges from 0 (no severity) to 61 (highest severity).

**Conclusion:** This study represents a first step towards the development of a multidimensional TB severity assessment tool for PLWH. However, its clinical usefulness, feasibility, and added value compared with existing severity scores remain to be demonstrated through validation studies before routine implementation can be considered.

## INTRODUCTION

Tuberculosis (TB) remains the leading cause of death in people living with the Human Immunodeficiency Virus (PLWH), accounting for 161,000 deaths in 2023, and is the most frequent opportunistic infection (OI) in this population, with 662,000 estimated TB disease in the same year.^1^ Beyond mortality, TB in PLWH is associated with substantial morbidity, including conditions requiring hospitalization or prolonged hospitalization, paradoxical clinical deterioration due to Immune Reconstitution Inflammatory Syndrome (IRIS), delayed cure, treatment failure or relapse.^2,3^ Better identifying PLWH with severe forms of TB could help target early interventions such as closer medical surveillance or differentiated treatment options depending on the level of severity to reduce mortality and severe morbidity.^4^

Multiple studies have assessed factors associated with mortality and different aspects of morbidity, i.e., severity in adults with TB living with or without HIV and several have proposed scoring systems for evaluation of severity.^7–14^ In a previously published systematic review, we identified 16 TB severity scores, including 5 (31.2%) specifically developed for PLWH with TB.^6^ Among main factors determining TB severity are characteristics related to the disease itself such as dissemination, extent of pulmonary involvement, and bacillary load.^3,5^ However, none of the existing TB severity assessment tools include all these factors.

The oldest severity score, the Bandim TBscore that predicts 8 months mortality in TB patients from low-resource settings, and its simplified and widely used version, the TBscore II that predicts 6 months mortality, are based solely on clinical criteria (cough, hemoptysis, dyspnea, chest pain, night sweating, anemia, tachycardia, abnormal lung auscultation, fever, low BMI and low mid-upper arm circumference (MUAC)).^7,8^ The Timika score and the Simplified Chest X-ray Score grades chest X-ray (CXR) severity and predict treatment outcomes in patients with smear-positive pulmonary TB based on the percentage of lung affected and presence of cavitation.^9,10^ The CABI score predicts mortality during TB treatment based on clinical form of TB, age, adjusted BMI, and HIV infection.^11^ The ICU tuberculosis score, the SCCOR-TB, and the score proposed by Singla et al. predict mortality in patients admitted in intensive care unit or emergency department based respectively on miliary tuberculosis, mechanical ventilation and vasopressive requirements; septic shock, HIV infection, serum creatinine, ratio of arterial partial pressure of oxygen by fractionated inspired oxygen, diffuse parenchymal infiltrates or a miliary pattern on CXR, and absence of antituberculosis treatment on admission; SpO₂, respiratory rate, systolic blood pressure, heart rate, and advanced disease on CXR. ^12–14^ The TB prognostic score assesses the prognosis of tuberculosis in smear positive HIV-negative patients based on age, respiratory failure requiring oxygen, serum albumin and functional status.^15^ The TB mortality risk score predicts mortality during treatment based on age, US born status, homelessness, residence in a long-term care facility, chronic kidney disease, meningeal tuberculosis, miliary tuberculosis, specific tuberculosis abnormality on CXR, and HIV status. ^16^ The Tuberculosis risk assessment tool (TReAT) stratifies 6-month mortality risk in patients with pulmonary TB based on hypoxemic respiratory failure, age, bilateral pulmonary involvement, haemoglobin, and significant comorbidity (HIV, diabetes mellitus, liver failure, cirrhosis, congestive heart failure, chronic respiratory disease). ^17^ Only the Bandim TBscore, its simplified version, the TBscore II, and the Timika score have been externally validated or used in other studies.^18,19^

Among the TB severity scores specifically developed for PLWH, the Health Care Index (HCI) and its updated version, the new Health Care Index, that predict 12-month mortality, are based on TB drug resistance, inclusion of rifamycin (R), isoniazid (H) and pyrazinamide (Z) in the initial TB treatment, antiretroviral therapy (ART), and viral load, parameters reflecting care provided to patients rather than specific characteristics of the disease.^20,21^ The Nguyen score, that predicts mortality during TB treatment in hospitalized patients based on age, residence in a long-term care facility, meningeal tuberculosis, radiological abnormalities consistent with TB, and microbiological status, was developed solely from data of patients in the United States and may therefore not be applicable in settings with a high HIV/tuberculosis prevalences.^22^ The Gupta-Wright score that predicts 2-months mortality based on age, sex, ongoing ART, anemia, inability to walk unaided and lipoarabinomannan (LAM) test result, does not include radiological criteria.^23^ At last, the clinical scoring model developed by Zhang *et al.* predicts mortality at 12 months of follow-up after hospitalization at AIDS end stage based on hemoglobin, tuberculous meningitis, severe pneumonia, hypoalbuminemia, unexplained infections or space-occupying lesions, and malignant tumors.^24^ However, it was shown that incidence of death is the highest at the beginning of hospitalization.^25–27^

^A^lthough they may be clinically useful, existing TB severity assessment tools may be sub-optimal for assessing disease severity in PLWH in high TB burden low resources settings, since i incompletely integrate TB-specific factors and very few were tailored to that population. In that context, we sought to develop a consensus-based multifactorial TB severity score specifically tailored to PLWH by combining the best judgement of a multidisciplinary panel of international TB experts with the evidence provided by a systematic review.

## METHODS

We implemented a *RAND/University of California Los Angeles Appropriateness Method* (RAM) process to design a severity scoring system for TB in PLHIV The RAM uses a modified Delphi panel approach to combine expert opinion with available evidence to assess appropriateness of practices in defined clinical situations. The available evidence is provided by a detailed literature review and the modified Delphi is usually composed of two rounds: round 1 consists of a questionnaire summarizing the indications provided by the literature review whose appropriateness is independently rated by a panel of experts; round 2 consists of a meeting during which experts discuss their ratings, focusing on areas of disagreement, and then re-rate each indication individually; no attempt is made to force the panel to consensus.^28^ The RAM protocol was posted on the PROSPERO platform (CRD42022323983). Our systematic review was published elsewhere.^6^ We report here on the modified Delphi process results.

### Conduct of the RAM process

We gathered an expert panel selected on the following criteria: i) at least 10 years of experience in the field of TB; ii) peer-recognized leadership in their specialty (infectious disease, microbiology, pulmonology, clinical immunology or epidemiology); iii) good command of the French language. We identified experts based on the recommendations of the International Union Against Tuberculosis and Lung Disease and of other experts. The experts did not receive any compensation for their participation.

Before the start of the RAM process, a first meeting was held with the panel to reach a consensus-based definition of severe TB: it was agreed that severe TB should present at least one of the following potential risks: death, severe morbidity or sequelae.

Following a systematic review that identified factors associated with TB severity from the literature in participants aged ≥15 years, we designed an online questionnaire aiming to assess the appropriateness of including those factors that were identified in a TB severity score. The questionnaire was developed on the Survey Monkey platform (surveymonkey.com). We then held a round zero meeting with the panel to present the RAM process, the results of the systematic review, the questionnaire and the definitions of the terms used.

After the round 0 meeting, experts were given 6 weeks to independently assess the TB severity factors via the online questionnaire based on both the systematic review report (which was sent to them in advance) and their best judgment (Round 1). They were given the opportunity to add comments and any other factor that they might think relevant to include in the severity score. For each factor, each expert was asked to: i) rate the appropriateness of including the factor in the TB severity score by giving it a score between 1 and 9, from not at all appropriate and entirely appropriate to include this factor in the score; and ii) grade the level of severity associated with the factor from low, to moderate, high or very high.

At round 2, two meetings of two hours each were conducted to discuss the results of the first round. Factors added by experts during the first round were also discussed. At the end of each meeting, the experts conducted a second assessment of severity factors using the online questionnaire within 24 hours. Both meetings were conducted by a highly experienced moderator with a good understanding of the RAM process.

### Consensus building

Each severity factor was classified into one of the following three levels of appropriateness, based on expert ratings of the appropriateness of including it in the severity score : i) high: median expert rating from 7 to 9, with no disagreement (meaning the number of experts rating 1 to 3 is not too high); ii) uncertain: median rating from 4 to 6 or any median with disagreement; iii) low: median rating from 1 to 3, with no disagreement (meaning the number of experts rating 7 to 9 is not too high). Only those severity factors with a high level of appropriateness were finally included in the severity score. The levels of disagreement were defined by the panel size and the number of panelists rating in each extreme (1-3 and 7-9). In a panel of 8 experts, there is disagreement when at least 3 vote at the other extreme of the median; in a panel of 14 experts, there is disagreement when at least 5 vote at the other extreme of the median.^28^ Each factor was given the severity level which reached >50% of votes. The severity level of the factors not reaching >50% of votes in any severity level was classified as uncertain.

### Statistical analysis

The data were entered into Microsoft Excel to calculate the ratings percentages, medians and levels of disagreement.

### Ethical considerations

All studies included in the systematic review reported being approved by ethics committees.

## RESULTS

The round 1 questionnaire was sent to 15 experts, among which 8 (53.3%) responded, including 3 infectious disease specialists, 1 microbiologist, 2 pulmonologists, 1 clinical immunologist and 1 epidemiologist from Benin (1), Burkina Faso (1), Côte d’Ivoire (1) and France (5). All those who responded attended both round 2 meetings.

Of 62 factors considered by experts for rating, 15 (24.2%) factors were considered as having a high level of appropriateness for the severity score, including: proportion of lung involvement, number of cavitations, urine LAM test results, extrapulmonary involvement, hemoptysis, dyspnea, respiratory rate, pulse oxygen saturation (SpO_2_), partial pressure of oxygen (PaO^2^), BMI, functional status, hemoglobin, CD4 count, number of concomitant opportunistic infections, comorbidities); 15 (24.2%) were considered uncertain and 32 (51.6%) with a low level (Table 1). Since the panel size was 8, disagreement was defined as a number of experts rating in each extreme (1-3 and 7-9) of ≥ 3. The detailed ratings of the appropriateness of including factors in the severity score are presented in Table S1.

**Table 1.** Appropriateness of including factors in the severity score.

| High (n=15) | Uncertain (n=15) | Low (n=32) |
| --- | --- | --- |
| Proportion of lung involvement | Clinical anemia | Chronic cough (> 2 weeks) |
| Number of cavitations | Heart rate | Chest pain |
| Urinary LAM | Systolic blood pressure | Anorexia |
| Extrapulmonary involvement | Number of previous tuberculosis episodes | Asthenia |
| Hemoptysis | Dehydration | Night sweating |
| Dyspnea | Delay in tuberculosis diagnosis | Temperature |
| Respiratory rate | Delay in treatment | Abnormal lung auscultation |
| Pulse Oxygen Saturation (SpO2) | WHO AIDS stage | Pregnancy |
| Partial Pressure of Oxygen (PaO2) | Age | Sex |
| Body Mass Index (BMI) | Drug abuse | Education |
| Functional status | Poverty | Residence |
| Hemoglobin | Microscopy result | Long-term care facility |
| CD4 count | Microscopy positivity grade | Prison |
| Number of concomitant opportunistic infections | Xpert MTB/RIF positivity level | Tobacco |
| Comorbidities | Immunosuppressive medications | Alcohol |
|  |  | Homeless |
|  |  | Marital status |
|  |  | No physical activity |
|  |  | Occupation |
|  |  | Traditional medicine/Herbal medicine |
|  |  | Displaced population |
|  |  | Culture result |
|  |  | Xpert MTB/RIF result |
|  |  | C-reactive protein (CRP) |
|  |  | Uremia |
|  |  | Blood glucose |
|  |  | Blood albumin |
|  |  | Blood sodium |
|  |  | Antiretroviral (ARV) treatment |
|  |  | No cotrimoxazole prophylaxis |
|  |  | No isoniazid prophylaxis |
|  |  | Negative tuberculin skin test |

In addition to factors included in the final severity score due to their high level of appropriateness, we included microscopy positivity grade as well, as it had a median rating of 6 (right below the high level of appropriateness) without disagreement, and 75% of the experts rated that a grade 3+ smear positive was associated with at least a moderate severity level. We chose SpO_2_ over PaO_2_ based on the experts’ advice and practicality of measurement. Only comorbidities or extrapulmonary involvement with very high or high severity level were kept in the score for practicality.

The severity levels associated with the factors included in the severity score are presented in Table 2. The detailed ratings are presented in Table S2. To build the multifactorial TB severity score (TBSS), numerical values were given to each severity level as follows: 0 for low, 1 for moderate, 2 for high, and 3 for very high. The sum of the numerical values yielded a total score ranging from 0 (no severity) to 61 (highest severity). There are 9 factors with moderate severity levels, yielding a maxiumu score of 9; There are 18 factors with high severity levels, yielding a maxiumu score of 36 ; based on this, the TBSS was split into four severity classes with a proposed management according to each severity class (Table 3).

**Table 2.** Severity levels associated with the factors included in the multifactorial TB severity score.

| Factors | Severity levels |  |  |  | Maximum score |
| --- | --- | --- | --- | --- | --- |
|  | Very high<br>(3) | High<br>(2) | Moderate<br>(1) | Low<br>(0) |  |
| Total proportion of lung involvement (%) | ≥70 | [30-70[ | [10-30[ | <10 | 3 |
| Number of cavitations |  | ≥3 | 1-2 | 0 | 2 |
| Urinary LAM | Positive |  |  | Negative | 3 |
| Extrapulmonary involvement |  |  |  |  |  |
| Meningeal/cerebral | Presence |  |  | Absence | 3 |
| Spinal | Presence |  |  | Absence | 3 |
| Disseminated tuberculosis |  | Presence |  | Absence | 2 |
| Miliary TB |  | Presence |  | Absence | 2 |
| Pericardial |  | Presence |  | Absence | 2 |
| Microscopy positivity grade |  |  | 3+ | <3+ | 1 |
| Hemoptysis |  | Presence |  | Absence | 2 |
| Dyspnea<sup>a</sup> |  | Presence |  | Absence | 2 |
| Respiratory rate (cycles per minute) |  | >30 | ]20-30] | ≤20 | 2 |
| Pulse Oxygen Saturation (SpO<sub>2</sub>) (%) | <80 |  | [80-95[ | ≥95 | 3 |
| Body Mass Index (BMI) (kg/m<sup>2</sup>) | <16 | [16-18,5[ |  | ≥18,5 | 3 |
| Functional Status | Bedridden | Unable to walk<br>without<br>assistance |  | Completely<br>independent /<br>Reduced activities<br>of daily living | 3 |
| Hemoglobin Level (g/dL) | <7 | [7-8[ | [8-10[ | ≥10 | 3 |
| CD4 Count (cellules/μL) | <50 | [50-200[ | [200-350[ | ≥350 | 3 |
| Number of concomitant opportunistic infections |  | ≥2 | 1 | 0 | 2 |
| Number of comorbidities | ≥3 | 2 | 1 | 0 | 3 |
| Type of comorbidities |  |  |  |  |  |
| Liver cirrhosis | Presence |  |  | Absence | 3 |
| Chronic renal failure (GFR <50<br>ml/min/1.73 m<sup>2</sup>) | Presence |  |  | Absence | 3 |
| Impairment of higher functions<sup>b</sup> |  | Presence |  | Absence | 2 |
| Heart failure |  | Presence |  | Absence | 2 |
| Cancer |  | Presence |  | Absence | 2 |
| Stroke |  | Presence |  | Absence | 2 |
| **TOTAL MAXIMUM SCORE** |  |  |  |  | **61** |

<sup>a</sup> Subjective sensation of breathing discomfort that consists of qualitatively distinct sensations that vary in intensity; <sup>b</sup> cognitive impairment or dementia.

**Table 3.** Severity classes of the Multifactorial Tuberculosis Severity Score.

| Severity class | Score Criteria | Proposed Management |
| --- | --- | --- |
| **Low** | **0** | No monitoring, standard treatment |
| **Moderate** | **1–9, but no factor rated as high or very high severity** | Outpatient monitoring, standard treatment |
| **High** | **10–36, but no factor rated as very high severity** | Close monitoring during hospitalization, consider treatment intensification |
| **Very high** | **>36 or presence of $\geq 1$ factor rated as very high severity** | Monitoring in intensive care unit, consider treatment intensification |

## DISCUSSION

Using the RAND/UCLA Appropriateness Method, we developed a consensus-based multifactorial TB severity score (TBSS) for PLWH. The score integrates 15 parameters encompassing TB-related characteristics (extent of lung involvement, bacillary load, and disease dissemination), host-related characteristics (nutritional status, immunosuppression, functional status, and comorbidities), and characteristics reflecting the interaction between the host and the disease. By combining these complementary dimensions, the TBSS aims to provide a comprehensive assessment of TB severity in PLWH.

A major finding of this study is the importance attributed by experts to key TB-related determinants of severity. The TBSS includes all three important TB-related characteristics that were previously reported to be associated with severity: bacillary load (microscopy positivity grade), extent of pulmonary involvement (total proportion of lung involvement and number of cavitations), and disease dissemination (positive urine LAM test and extrapulmonary involvement).^3,29–31^ To our knowledge, no existing severity score specifically developed for PLWH simultaneously captures all these dimensions.^20–24^ Their inclusion reflects growing evidence that disease burden is a central component of TB severity, particularly in immunocompromised populations.

The TBSS also incorporates several parameters that are absent from most existing scores, including SpO_2_, hemoptysis, CD4 count, number of concomitant OIs, BMI, dyspnea, respiratory rate, and hemoglobin which were reported to be associated with TB severity.^3,12,17,32^ The inclusion of these variables acknowledges that TB severity in PLWH is not determined solely by characteristics of the infection itself but also by the host’s capacity to respond to disease. For example, severe anaemia, advanced immunosuppression, or profound malnutrition may substantially worsen prognosis independently of mycobacterial burden. Conversely, apparently moderate TB disease may result in poor outcomes in patients with severe immune deficiency. The final score therefore reflects the multifactorial nature of TB severity in PLWH and the complex interplay between pathogen-related and host-related determinants.

Our study should be interpreted in light of several limitations. First, the response rate in the first Delphi round was 53%, which may raise concerns regarding representativeness and potential selection bias. However, all participating experts completed both Delphi rounds, and the panel included specialists from multiple disciplines and from both high- and low-burden HIV/TB settings. In addition, the panel size exceeded the minimum number recommended in the RAND/UCLA Appropriateness Method manual.^28^ Second, the TBSS includes 15 parameters, which may limit its feasibility in some clinical settings, particularly where access to laboratory investigations or imaging is restricted. Although most variables are routinely collected during HIV/TB care, future studies should evaluate feasibility and determine whether simplified versions of the score could retain adequate performance while improving usability. Nevertheless, the number of parameters included in the TBSS remains comparable to that of other TB severity scores and widely used clinical severity indices. The Bandim TBscore has 11 parameters, the acute physiology and chronic health evaluation (APACHE) has 18, and the simplified acute physiology score (SAPS) has 21.^7,33,34^ Third, the score has not yet undergone formal validation. Consequently, its measurement properties, predictive performance, responsiveness to clinical change, and operational utility remain to be established.

Despite these limitations, our study has some strengths. The TBSS was developed using a rigorous and transparent methodology that combined evidence from a previously published systematic review with the collective judgement of an international multidisciplinary expert panel. This approach ensured that the score was grounded in both empirical evidence and clinical experience. Furthermore, the score was specifically designed for PLWH, a population in whom TB presentations are frequently heterogeneous and strongly influenced by the degree of immunosuppression. By integrating TB-related, host-related, and mixed determinants of severity, the TBSS provides a structured framework that reflects the complexity of TB disease in that population..

Further work is now required to evaluate the performance of the TBSS in clinical datasets and prospective cohorts. Future studies should assess its content validity, construct validity, criterion validity, responsiveness to change, feasibility, and predictive performance for clinically relevant outcomes such as mortality, and severe morbidity. Comparative studies against existing tools, including the TBscore II, will also be necessary to determine whether the TBSS provides additional prognostic or clinical value.

## CONCLUSION

This study represents a first step towards the development of a multidimensional TB severity assessment tool for PLWH. Developed through a structured consensus process informed by a systematic review, the TBSS integrates parameters encompassing TB-related characteristics, host-related characteristics, and characteristics reflecting the interaction between the host and the disease. However, its clinical usefulness, feasibility, and added value compared with existing severity scores remain to be demonstrated through validation studies before routine implementation can be considered.

**Figure 1.**
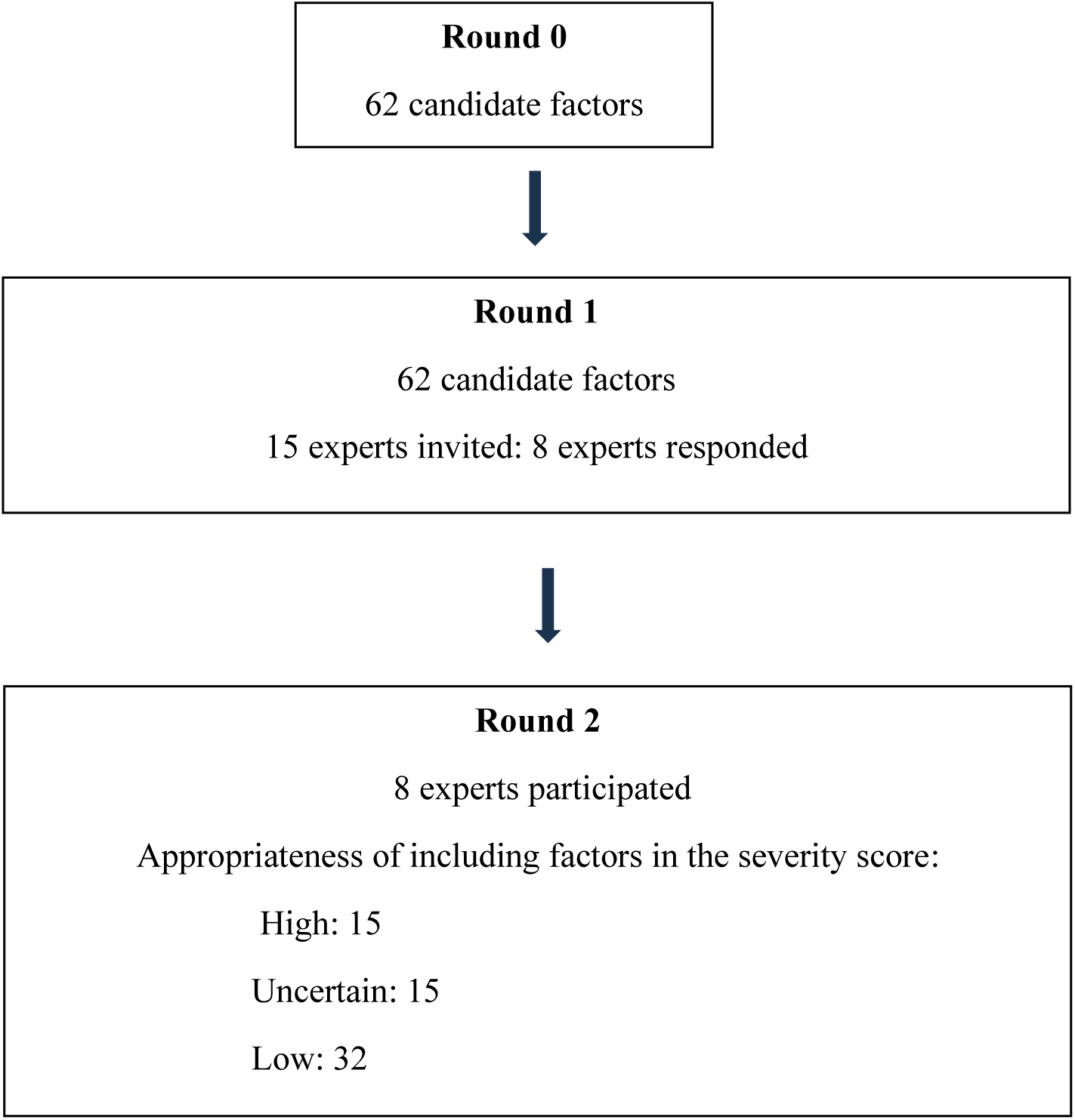
Selection of factors for the tuberculosis severity score using the RAND/UCLA Appropriateness Method

## Data Availability

All data produced in the present study are available upon reasonable request to the authors

## ACKNOWLEDGEMENTS

The authors have not declared a specific grant for this research from any funding agency in the public, commercial or not-for-profit sectors.

Author contributions: RA was in charge of study conception and design under the supervision of MZ and OM, developed data collection tool, did the analyses and drafted the manuscript. OM revised the first draft. DL, CAT, NDC, GB, NV, EM, FXB and LW were members of the expert panel. XA was the moderator of the meetings. All authors revised the manuscript for intellectual content and approved the final version. All authors have read and agreed to the submitted version of the manuscript.

---

**Supplementary Table S1.** CHARMS checklist applied to the development of the TBSS.

| **Domain** | **Item** | **Description in this study** |
| --- | --- | --- |
| **Source of data** | Study design | Development of a severity score using the RAND/UCLA Appropriateness Method based on a prior systematic review |
|  | Data source | Published literature (systematic review) and expert opinion (Delphi panel) |
| **Participants** | Study population | Not applicable (no patient-level data); expert panel included clinicians and researchers with ≥10 years of experience in TB |
|  | Setting | Experts from high- and low-burden TB/HIV settings (Africa and Europe) |
|  | Inclusion criteria | Experts with recognized expertise in TB (infectious diseases, pulmonology, microbiology, immunology, epidemiology) |
| **Outcome to be predicted** | Outcome definition | TB severity, defined as risk of combined death, severe morbidity |
|  | Outcome measurement | Consensus-based definition established prior to Delphi rounds |
| **Candidate predictors** | Predictor definition | Factors associated with TB severity identified through systematic review |
|  | Number of predictors | 62 candidate factors evaluated |
|  | Measurement of predictors | Based on literature definitions and expert interpretation |
| **Sample size** | Number of participants | 15 experts invited; 8 participated in round 1 and 2 |
| **Missing data** | Handling | Not applicable (expert ratings only; no missing ratings among respondents) |
| **Model development** | Model type | Consensus-based scoring system (not statistical model) |
|  | Predictor selection | Based on appropriateness ratings (median 7–9 without disagreement) |
|  | Weighting of predictors | Assigned based on severity levels (0–3) determined by expert consensus |
| **Model performance** | Performance measures | Not assessed (no validation dataset) |
|  | Internal validation | Not performed |
|  | External validation | Not performed |
| **Results** | Final model | TB severity score including 15 parameters |
|  | Score range | 0 to 61 |
| **Interpretation** | Clinical use | Proposed tool to structure assessment of TB severity in PLWH |
| **Limitations** | Key limitations | Small expert panel, 53% response rate, lack of validation, feasibility concerns |
| **Funding** | Funding source | No specific funding declared |

